# Value of information dynamics in Disease X vaccine clinical trials

**DOI:** 10.1101/2023.05.26.23290603

**Authors:** Nicolas Houy, Julien Flaig

## Abstract

**Background:** Solutions have been proposed to accelerate the development and rollout of vaccines against a hypothetical disease with epidemic or pandemic potential called Disease X. This may involve resolving uncertainties regarding the disease and the new vaccine. However the value for public health of collecting this information will depend on the time needed to perform research, but also on the time needed to produce vaccine doses. We explore this interplay, and its effect on the decision on whether or not to perform research.

**Method:** We simulate numerically the emergence and transmission of a disease in a population using a susceptible-infected-recovered (SIR) compartmental model with vaccination. Uncertainties regarding the disease and the vaccine are represented by parameter prior distributions. We vary the date at which vaccine doses are available, and the date at which information about parameters becomes available. We use the expected value of perfect information (EVPI) and the expected value of partially perfect information (EVPPI) to measure the value of information.

**Results:** As expected, information has less or no value if it comes too late, or (equivalently) if it can only be used too late. However we also find non trivial dynamics for shorter durations of vaccine development. In this parameter area, it can be optimal to implement vaccination without waiting for information depending on the respective durations of dose production and of clinical research.

**Conclusion:** We illustrate the value of information dynamics in a Disease X outbreak scenario, and present a general approach to properly take into account uncertainties and transmission dynamics when planning clinical research in this scenario. Our method is based on numerical simulation and allows us to highlight non trivial effects that cannot otherwise be investigated.

## 1 Introduction

A host of infectious diseases – the Spanish influenza, Ebola, COVID-19, to name a few – were unknown to scientists or subject to substantial uncertainties when they emerged as epidemic or pandemic threats [1]. In 2018, WHO introduced the placeholder “Pathogen X” referring to an infectious agent with epidemic or pandemic potential that is not currently known to cause human illness. The disease caused by Pathogen X was dubbed “Disease X” [2]. The objective of investigating hypothetical disease Disease X is to prepare against the emergence of yet unknown pathogens and accelerate the development of medical countermeasures in case of outbreak. More recently, the Coalition for Epidemic Preparedness Innovations (CEPI) set the challenge goal of being able to produce and distribute a vaccine at scale within 100 days after a potentially pandemic pathogen is detected [3].

Technological, organizational, and regulatory solutions have been proposed to shorten vaccine development in the event of a Disease X outbreak [3–5]. For instance, libraries of immunization targets, adjuvants, or rapid-response vaccine platforms, as well as appropriate manufacturing approaches, could be developed and validated in advance for known pathogens or broad families of pathogens. Similarly, clinical trial protocols could be designed and approved in advance – suggested protocols would, for example, merge clinical phases 1 and 2 or include rolling review of evidence.

In summary, these solutions consist in (i) accelerating the process of making vaccine doses physically available, which involves detecting the pathogen, developing vaccine candidates and producing doses at scale, and (ii) accelerating clinical trials to inform the decision on whether and how to use a vaccine candidate based on its estimated safety and efficacy. In a Disease X outbreak scenario, acceleration strategies of vaccine production and clinical research are meant to be implemented in conjunction with emergency vaccination of target risk groups [3]. This clearly raises the issue of balancing collection of clinical evidence and swift vaccine rollout. Generally, resolving uncertainties regarding a vaccine or an emerging disease can help control the outbreak more efficiently, but collecting information also takes time and there is a risk that the epidemic gets out of hand [6]. Then, how do the costs and benefits of a clinical trial – and more generally the value of collecting information about Disease X – change as functions of (i) the time needed to produce vaccine doses, (ii) the time needed to run the trial or collect information about the disease, and (iii) the overall vaccine development time? In this respect, what is the interplay between the acceleration of dose production and that of clinical research?

These questions can be addressed through value of information analyses or clinical trials design methods. Value of information analyses quantify the additional value of a clinical trial given current uncertainties, while trial design determines a specific trial implementation that meets an objective (e.g. a sample size that satisfies a given statistical power conditionally on a treatment effect). Both are closely related since value of information analyses provide upper bounds on the cost of clinical trial designs. Early studies in these areas typically focused on choosing a sample size to maximize utility, minimize costs, or equivalent [7–14]. Interestingly, several of these studies properly took uncertainty – a defining feature of Disease X – into account by considering parameter prior distributions [8–10, 12, 14]. However they were not specific to infectious disease countermeasures and most often sought to provide analytical results in static (not depending on time) settings. This, of course, is inappropriate in emerging infectious disease scenarios [15]. The dynamics of disease transmission proved particularly critical in designing clinical trials during the 2013–2016 Ebola crisis, because the epidemic was already waning when vaccines were ready for testing, which made sampling challenging [16, 17]. There is a rich literature using numerical simulations of dynamic disease transmission models to design Ebola vaccine clinical trials [18–22] – see also [23–25] for other diseases. While these studies take the dynamic of disease transmission into account, they mostly ignore uncertainties regarding the disease and countermeasures, or they treat them somewhat superficially by considering a range of scenarios. True treatment effects, in particular, are typically assumed. Besides, this literature usually takes the perspective of a statistician rather than a public health perspective: the performance of a clinical trial is measured in terms, e.g, of its statistical power rather than in terms of averted costs or disutility, that is in terms of the full value of the trial ([23] is an exception in this respect).

In this article, we illustrate the effect of the duration of vaccine development, that is the time needed to produce doses and to conduct clinical research, on the public health value of a clinical trial in a Disease X outbreak scenario. We consider both the epidemic dynamics, and uncertainties regarding Disease X *and* the vaccine in our reasoning. We model the spread of Disease X using a susceptible–infectious–recovered (SIR) model with vaccination. The disease parameters are assumed to be uncertain, as well as vaccine efficacy and safety – see [26, 27] for examples of adverse events discovered only at a late stage of vaccine development. Both infection and vaccine adverse effects have a cost and our goal is to minimize the total cost at the population level over the course of the epidemic. Clinical research is seen as a way to collect information about the disease and the vaccine to implement vaccination optimally. We use the expected value of perfect information (EVPI, see [28]) to estimate the value of entirely resolving all uncertainties, and the expected value of partial perfect information (EVPPI) to estimate the value of resolving uncertainties regarding subsets of parameters, in particular vaccine parameters that would typically be investigated by a clinical trial. We estimate EVPI and EVPPI for different durations of vaccine development. This value of information approach only provides upper bounds on the cost of clinical research. While actual costs are important in practice, they are highly context-specific. We believe that investigating upper bounds and their dynamics (as a function of development duration) is relevant for Disease X preparedness. Notice also that the reasoning presented in this article applies to any medical countermeasure and not only to vaccines.

Our method is detailed in Section 2: the transmission model in Section 2.1, and model uncertainties in Section 2.2. In Section 2.3, we formally derive an expression of EVPI and EVPPI in our scenario. The simulation results are gathered in Section 3. We start with an illustrative example of the trade-off between vaccinating under uncertainty and waiting for clinical research outcomes, and we provide cost breakdowns for this example (Section 3.1). Then we show simulations of decision making under uncertainty and perfect information (Sections 3.2 and 3.3 respectively). Finally, Section 3.4 gives the dynamics of EVPI and EVPPI as a function of the duration of clinical research and of vaccine dose production. Section 4 concludes.

## 2 Materials and methods

### 2.1 Disease transmission model

We consider the emergence of Disease X in a closed homogeneous population of *N* = 10^8^ individuals. At time 0, a single individual is infectious and the population is unvaccinated. We assume that the epidemic can be described by a deterministic SIR model with basic reproduction number *R*_0_ and mean duration of infectiousness 1*/γ*.

At time *T_v_*, a vaccine is available for rollout. Only susceptible individuals are vaccinated. Vaccination is successful with probability *p* – in our case, this parameter corresponds to vaccine efficacy and is equivalent to vaccine effectiveness. Successfully vaccinated individuals are immediately and fully protected, and there is no waning of vaccine protection. Independently of vaccination success, vaccinated individuals develop adverse events with probability *E*. The delay between vaccination and adverse events is exponentially distributed with mean *δ*. Figure App-1 shows a sketch of the transmission model.

We denote the cost per infectious day by *c_i_*, and the cost per adverse event by *c_a_*. *c_v_*is the cost of one administered vaccine dose, which includes production and operational costs. We ignore fixed costs for concision, and because we can plausibly assume that these costs have already been incurred at time 0 in our scenario.

### 2.2 Uncertainties

The model structure as well as the vaccine mechanism (all-or-nothing without loss of protection) are assumed to be known. This assumption can easily be relaxed by extending the model as appropriate. At time 0, transmission parameters, vaccine parameters, and costs are uncertain. This uncertainty is represented by parameter prior distributions. For the sake of illustration and discussion, we picked prior distributions yielding a range of results of interest while at the same time assuming plausible values.^1^ Figure 1 shows 10,000 draws from the parameter prior distributions. Disease transmission parameters *R*_0_ and *γ* are biologically plausible. The prior distributions of *δ* and *E* are assumed. We also assume low prior information regarding the efficacy parameter *p*. The order of magnitude of the cost *c_a_* of an adverse event broadly corresponds to that of damage payments, with the lower range corresponding to mild adverse events without compensation [29, 30]. The cost *c_i_* of an infectious days is of the order of magnitude of daily wages in high-income countries [31]. For the order of magnitude of the cost of a vaccine dose, see e.g. [32, 33]. We assume no parameter correlations.

**Figure 1:**
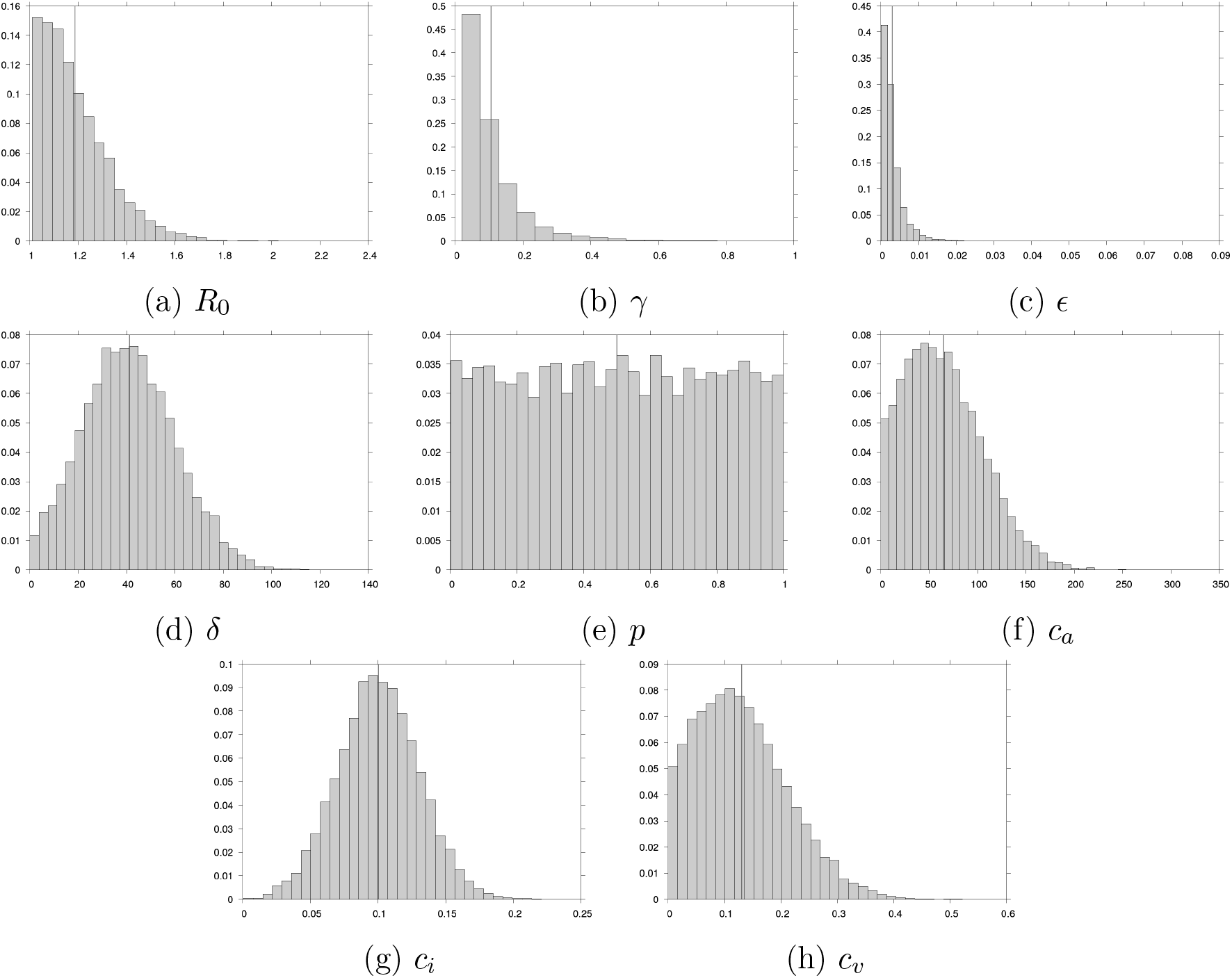
Frequency distributions for 10,000 draws of the parameters. Vertical lines show average values. The unit of time is one day. Costs are in thousand USD.

We assume that the true value of parameters can be known at time *T_v_* + *T_c_ ≥* 0. *T_c_ >* 0 means that this information is available after vaccine doses are available for rollout (at time *T_v_*), and *T_c_ <* 0 means that relevant information is available before completion of dose production. We are interested in the relationship between *T_v_*, *T_c_*, and the value of information brought by clinical research. We will therefore run simulations and estimate the value of information for different values of *T_v_* and *T_c_*, assuming both to be known. Notice that considering uncertain *T_v_* would only make decisions conditional on this value, whereas considering uncertain *T_c_* would not add any technical complexity while adding complexity for the sake of interpretation of our results.

### 2.3 Optimization problem and value of information

Our objective is to minimize the total cost over the course of the epidemic by deciding whether and when to vaccinate the population. For concision, we consider possible vaccination dates by increments of 30 days, and we assume that the whole susceptible population is vaccinated at once if we decide to vaccinate. These assumptions allow us to perform optimizations by exhaustively running all possible strategies when necessary.^2^ Let *A*(*t*) be the set of available vaccination policies consisting in vaccinating after date *t* or never vaccinating, and *C*(*a, ξ*) the total cost of policy *a* given parameter values *ξ*. Under uncertainty, a policy *a* is picked that minimizes the expected cost over parameter prior distributions. In this case, the set of available policies is *A*(*T_v_*) since vaccination can only be implemented after doses are available at time *T_v_*. The expected cost under uncertainty is then

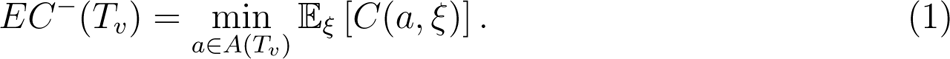

If parameter values are known at time *T_v_* + *T_c_*, a policy minimizing the cost *given these values* can be picked. In our scenario, if we assume that information can only be used once both vaccine doses and information are available, the set of available informed policies is *A*(max(*T_v_, T_v_* + *T_c_*)). In practice, however, decision making in the case *T_c_ <* 0 is very context-dependent – e.g. is it possible to stop vaccine production at *T_c_* depending on available information? – so we will focus on the case *T_c_≥* 0 and consider the set of available informed policies *A*(*T_v_* + *T_c_*). The expected cost of informed decision making is then

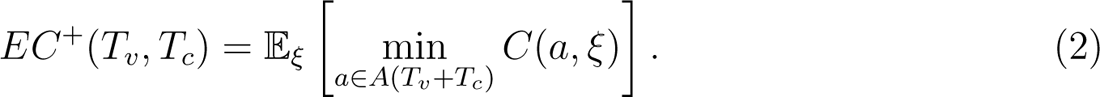

The difference *EC^−^*(*T_v_*)*− EC*^+^(*T_v_, T_c_*) is the expected value of using information about parameter values. Clearly, this value can be negative. If the expected cost of waiting for information is greater than that of vaccinating earlier under uncertainty, information is ignored.^3^ Thus the expected value of perfect information is in our case

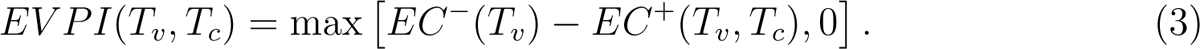

Clinical studies typically investigate only subsets of parameters (e.g. vaccine parameters in the case of vaccine clinical trials). The expected cost of choosing a policy knowing the true values for parameter subset *θ* is

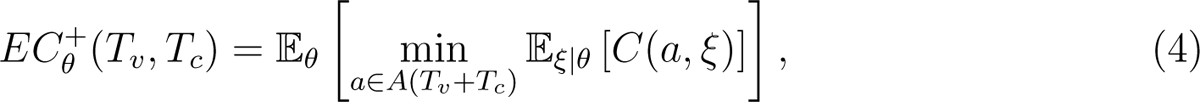

and the expected value of partially perfect information for *θ* is

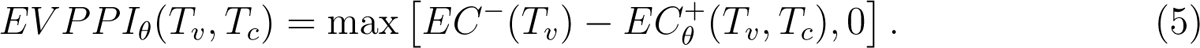

## 3 Results

### 3.1 An illustrative example

Table 1 and Figure 2 show the expected cumulative total cost over the course of the epidemic for three vaccination policies – no vaccination, vaccination at date 300, and vaccination at date 900 – and the cumulative total cost when parameters are set at their average value. Under uncertainty, in our scenario, vaccinating at date 300 would therefore be picked out of these three policies based on expected costs. With vaccination at date 900, the expected costs of vaccination and adverse events are lower compared to vaccination at time 300 because less individuals are still susceptible and eligible to vaccination by that time. Yet the expected overall cost of vaccinating at 900 is close to that of not vaccinating at all, the late policy implementation implying a higher number of infected individuals. Notice that if the vaccination decision was based on computations with parameters at their average value, vaccination would not be implemented.

**Figure 2:**
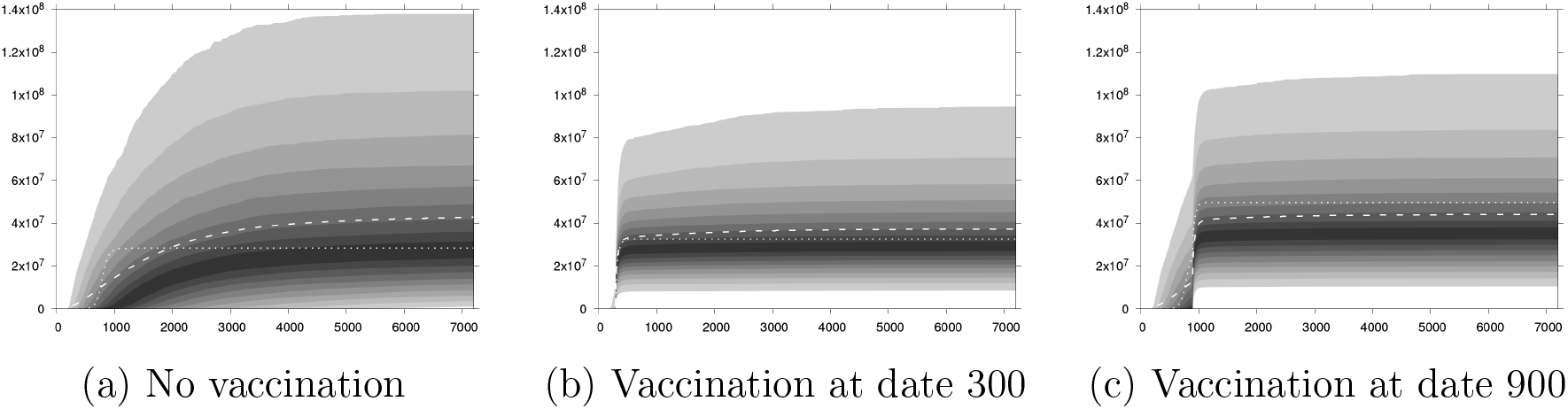
Cumulative total costs (thousand USD) as a function of time. Grey-black area for percentiles of simulations (10,000 draws from the prior distributions). Dashed line: expected cost over all simulations. Dotted line: simulation with parameters at their average values.

**Table 1:** Cumulative total costs (billion USD) for three vaccination policies. Expected costs are estimated over 10,000 draws from the prior distributions. 95% CIs are shown in parentheses.

|  | No vaccination | Vaccination<br>at date 300 | Vaccination<br>at date 900 |
| --- | --- | --- | --- |
| *Expected costs over prior distributions* |  |  |  |
| Infections | 44.02<br>(43.11 – 44.94) | 6.09<br>(5.73 – 6.45) | 16.09<br>(15.53 – 16.65) |
| Doses | 0<br>– | 12.55<br>(12.39 – 12.70) | 11.21<br>(11.06 – 11.35) |
| Adverse events | 0<br>– | 18.81<br>(18.34 – 19.29) | 16.86<br>(16.42 – 17.29) |
| Total | 44.02<br>(43.11 – 44.94) | 37.45<br>(36.84 – 38.07) | 44.15<br>(43.48 – 44.82) |
| *Costs for average parameter values* |  |  |  |
| Infections | 28.24 | $2.82 \times 10^{-3}$ | 25.83 |
| Doses | 0 | 12.99 | 9.50 |
| Adverse events | 0 | 19.59 | 14.33 |
| Total | 28.24 | 32.58 | 49.66 |

Let us now consider a scenario where vaccine doses are available at date 300 (*T_v_* = 300), and a hypothetical research program lets us know the exact value of all parameters at date 900 (*T_v_* + *T_c_* = 900). Under uncertainty for *T_v_*= 300, the best policy is to vaccinate at date 300 (see Figure 4 *infra*). Should we rather vaccinate at date 300 or wait and make a decision at date 900 knowing the true parameter values? Not vaccinating until date 900 and making a decision at date 900 knowing the true parameter values leads to vaccinating at date 900 for 32.88% of parameter values (95% CI: 31.96% – 33.80%) and not vaccinating for other parameter values. The expected cost of informed decision at date 900 is USD 29.03 billion (95% CI: 28.47 – 29.60), to be compared with the expected cost of decision under uncertainty at date 300 (Table 1): waiting for information leads to a gain of USD 8.42 billion (95% CI: 7.84 – 9.01). This is the maximum amount one should be willing to allocate to the considered research program.

Now, if information is made available at date *T_v_* + *T_c_* = 3000 instead of 900, the best policy at *T_v_* +*T_c_* is to vaccinate immediately for 9.86% of parameter values (95% CI: 9.28% – 10.44%) and to not vaccinate for other parameter values. The resulting expected cost of informed decision is USD 41.04 billion (95% CI: 40.16 – 41.93), which corresponds to an expected *loss* of USD 3.59 billion (95% CI: 2.65 – 4.52) compared to vaccinating at date *T_v_* = 300. With *T_v_* + *T_c_*= 3000, information would come too late so resources should not be allocated to the considered research program. Figure 3 shows the distributions of costs over parameter values when vaccination is implemented at date *T_v_* = 300 (uninformed decision), and when an informed decision is made at date *T_v_* +*T_c_* = 900 and *T_v_* +*T_c_* = 3000.

**Figure 3:**
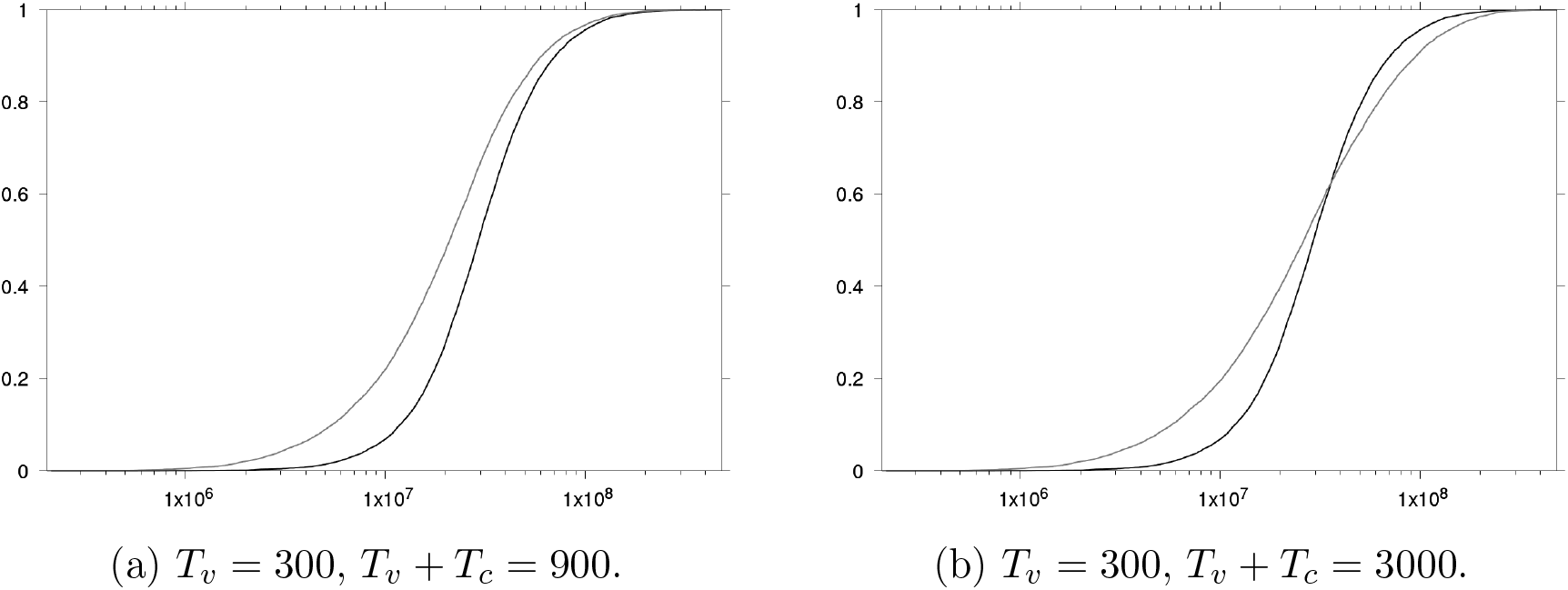
Cumulative distribution of total costs (thousand USD) for different values of *T_v_* and *T_v_* + *T_c_*. Black line: vaccination at *T_v_*. Grey line: informed decision at *T_v_* + *T_c_*.

### 3.2 Without information

Let us look into uninformed decision making in more details. In Figure 4, we display the expected cost difference between vaccinating at date *t* and not vaccinating at all as a function of *t*. For small vaccination dates, the epidemic is still at an early stage and the best policy is to vaccinate. As the epidemic spreads, the advantage of vaccination decreases and it becomes optimal not to vaccinate (under uncertainty). Formally, if we denote by *a_t_* the policy consisting in vaccinating at time *t* and by *a_∞_* the no vaccination policy, Figure 4 shows E*_ξ_* [*C*(*a_t_, ξ*) *− C*(*a_∞_, ξ*)] as a function of *t*. If vaccine doses are only available at time *T_v_*, we decide whether to vaccinate and at what date by considering the points such that *t ≥ T_v_*.

**Figure 4:**
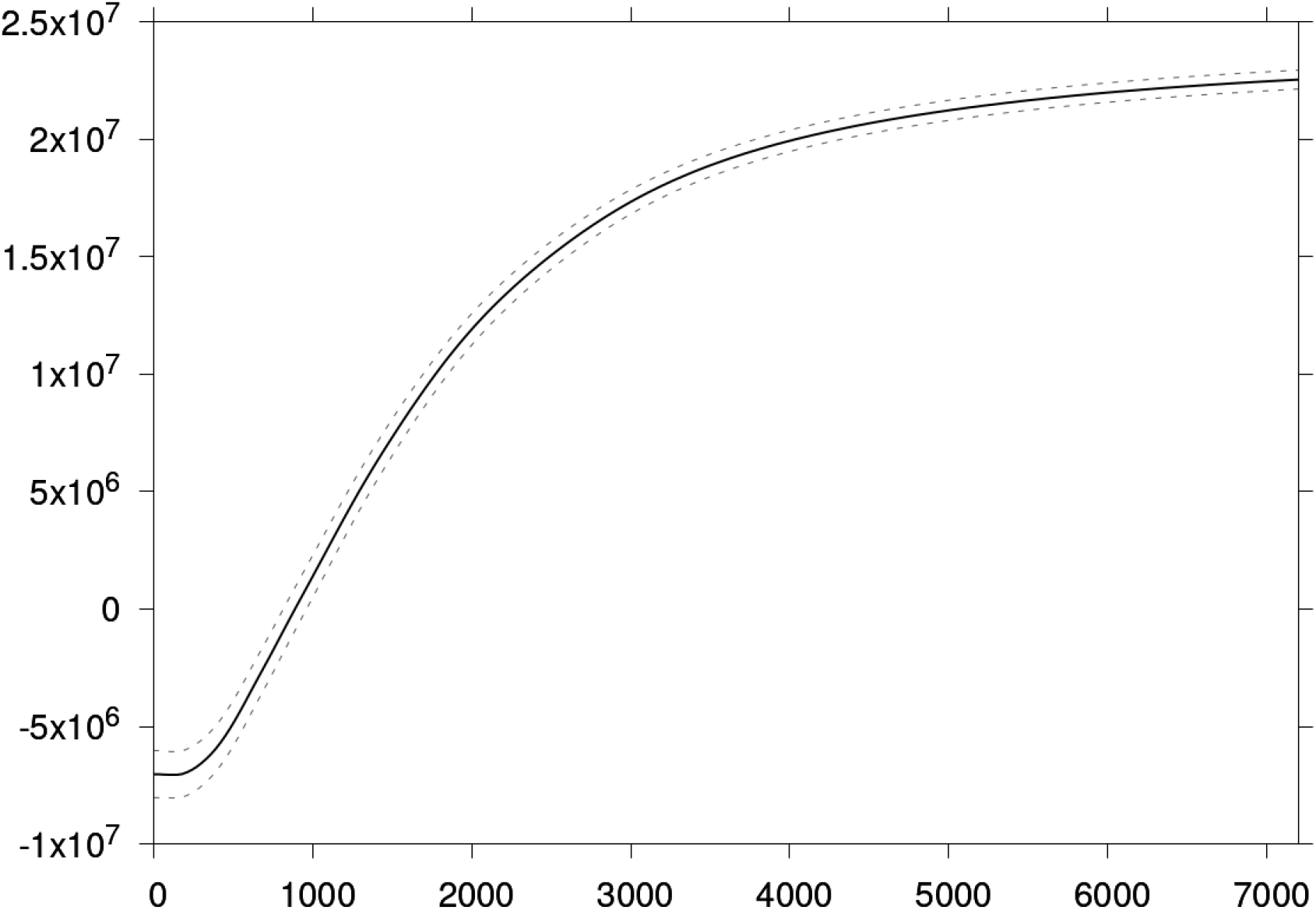
Expected cost difference (thousand USD) between vaccinating at date *t* and not vaccinating estimated over 10,000 parameter draws. x-axis: *t*. Dashed line: 95% CI.

### 3.3 With information

Now, let us assume that both the vaccine and true parameter values are available at date *T_v_* + *T_c_* and that the population is not yet vaccinated. At date *T_v_* + *T_c_*, we can then decide whether to vaccinate and at what date *t ≥ T_v_* + *T_c_* based on parameter values. Figure 5 shows the proportion of parameter values for which no vaccination is the best policy as a function of *T_v_* + *T_c_*. Again, vaccination becomes less advantageous as the epidemic is allowed to spread in our scenario. In Figure 6, we show the difference between the expected cost of making an informed decision, formally *EC*^+^(*T_v_, T_c_*), and the expected cost of not vaccinating, formally E*_ξ_* [*C*(*a_∞_, ξ*)]. For all *T_v_* +*T_c_*, making an informed decision is expected to be a better option than arbitrarily not vaccinating but the difference decreases over time.

**Figure 5:**
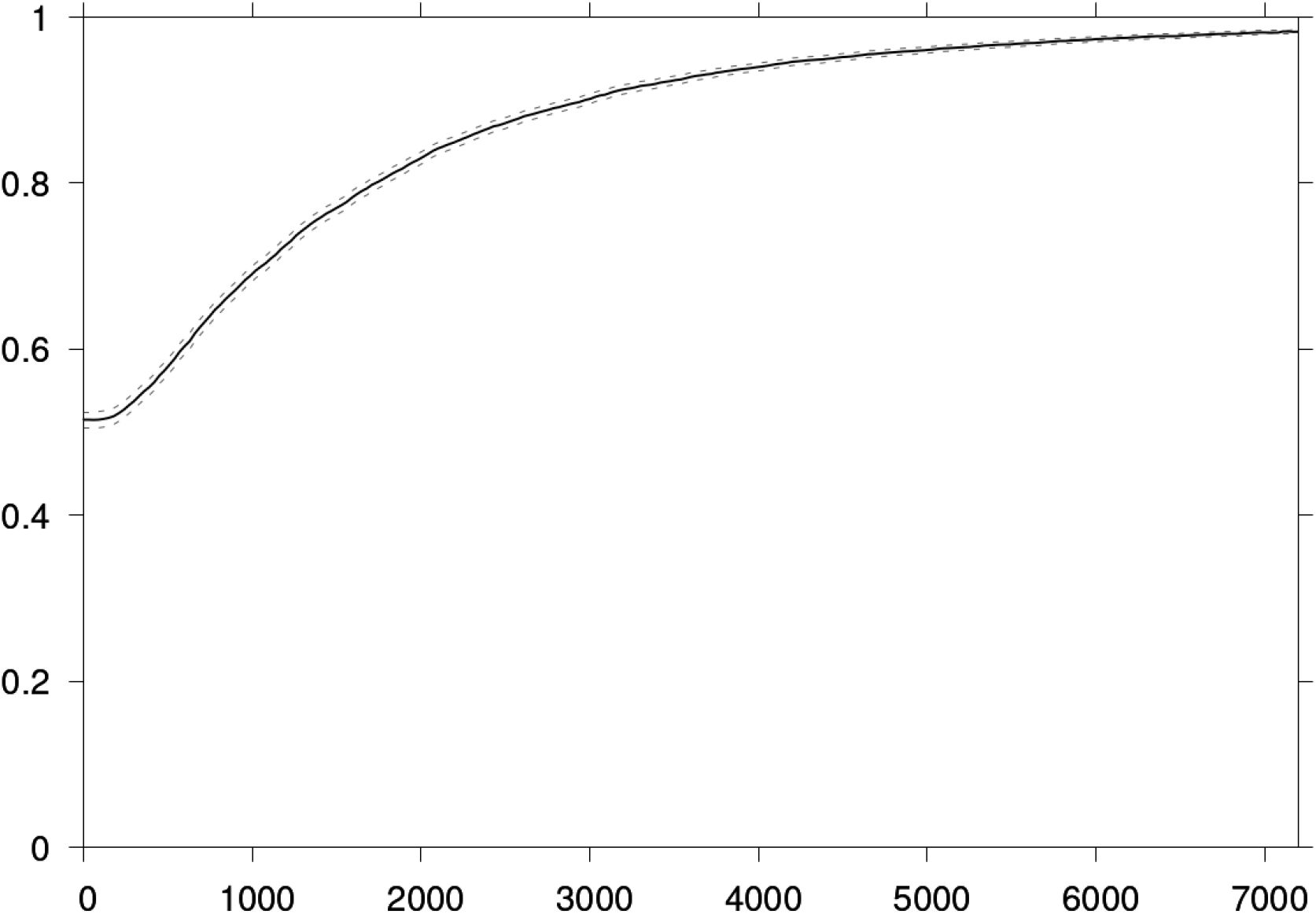
Expected proportion of cases for which no vaccination is implemented when information is available at date *T_v_* + *T_c_* and the decision maker has not yet vaccinated the population as a function of *T_v_* + *T_c_*. Estimation over 10,000 parameter draws. Dashed lines: 95% CI.

**Figure 6:**
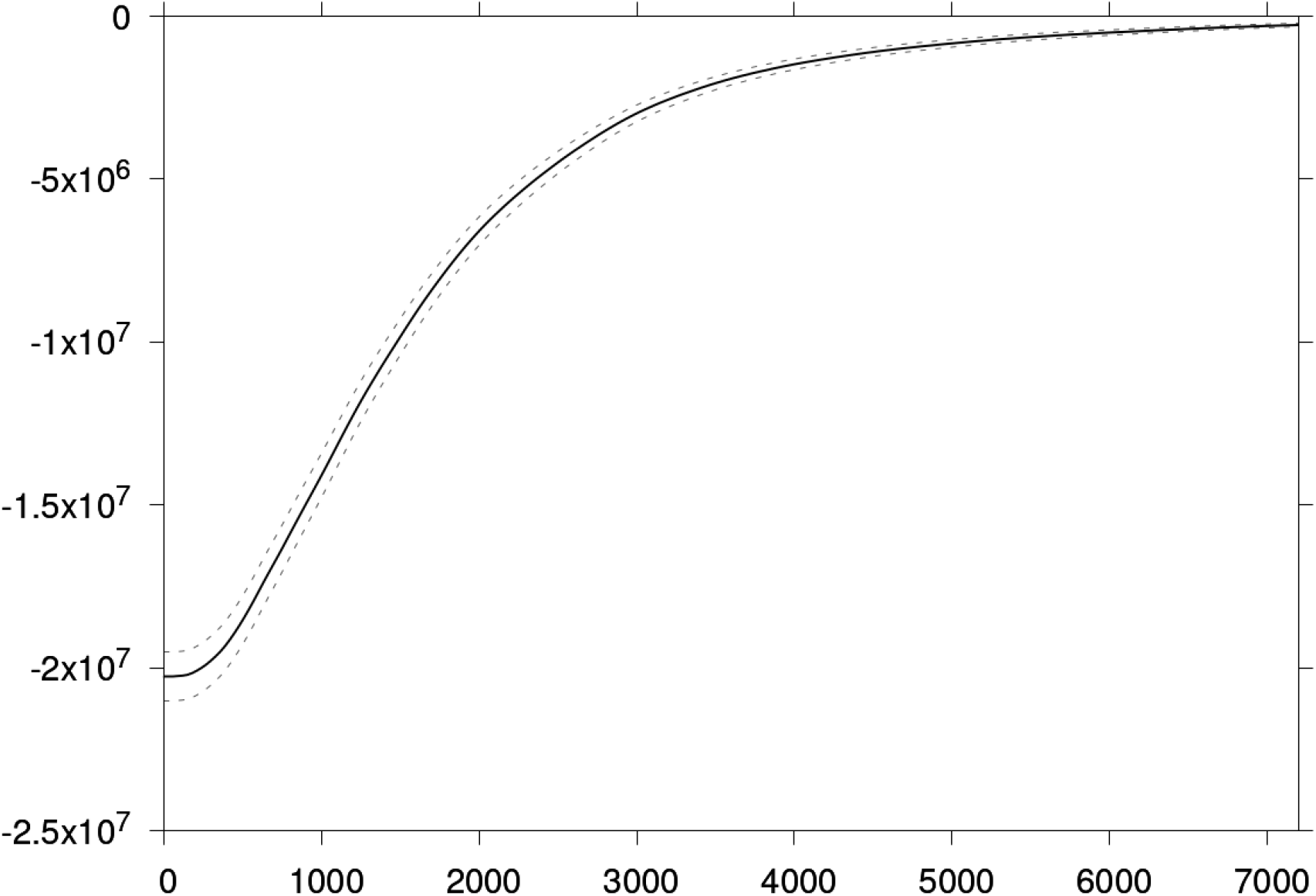
Expected difference (thousand USD) between the cost of the strategy consisting in vaccinating with information at date *T_v_*+*T_c_* and the strategy consisting in not vaccinating as a function of *T_v_* + *T_c_*. Estimation over 10,000 parameter draws. Dashed lines: 95% CI.

### 3.4 Value of information

From Equations 3 and 5 the value of information depends on both the time *T_v_* from which vaccine doses are available *and* the time *T_v_* + *T_c_*from which information is available. The EVPI dynamics is illustrated in Figure 7, and Figure 8 shows sectional views of the heatmap in Figure 7. A vaccine clinical trial would focus on vaccine parameters. In Figure 9, we display EVPPI for vaccine parameters. See Appendix D for EVPPI of other subsets of parameters. EVPPI was estimated following [34].

**Figure 7:**
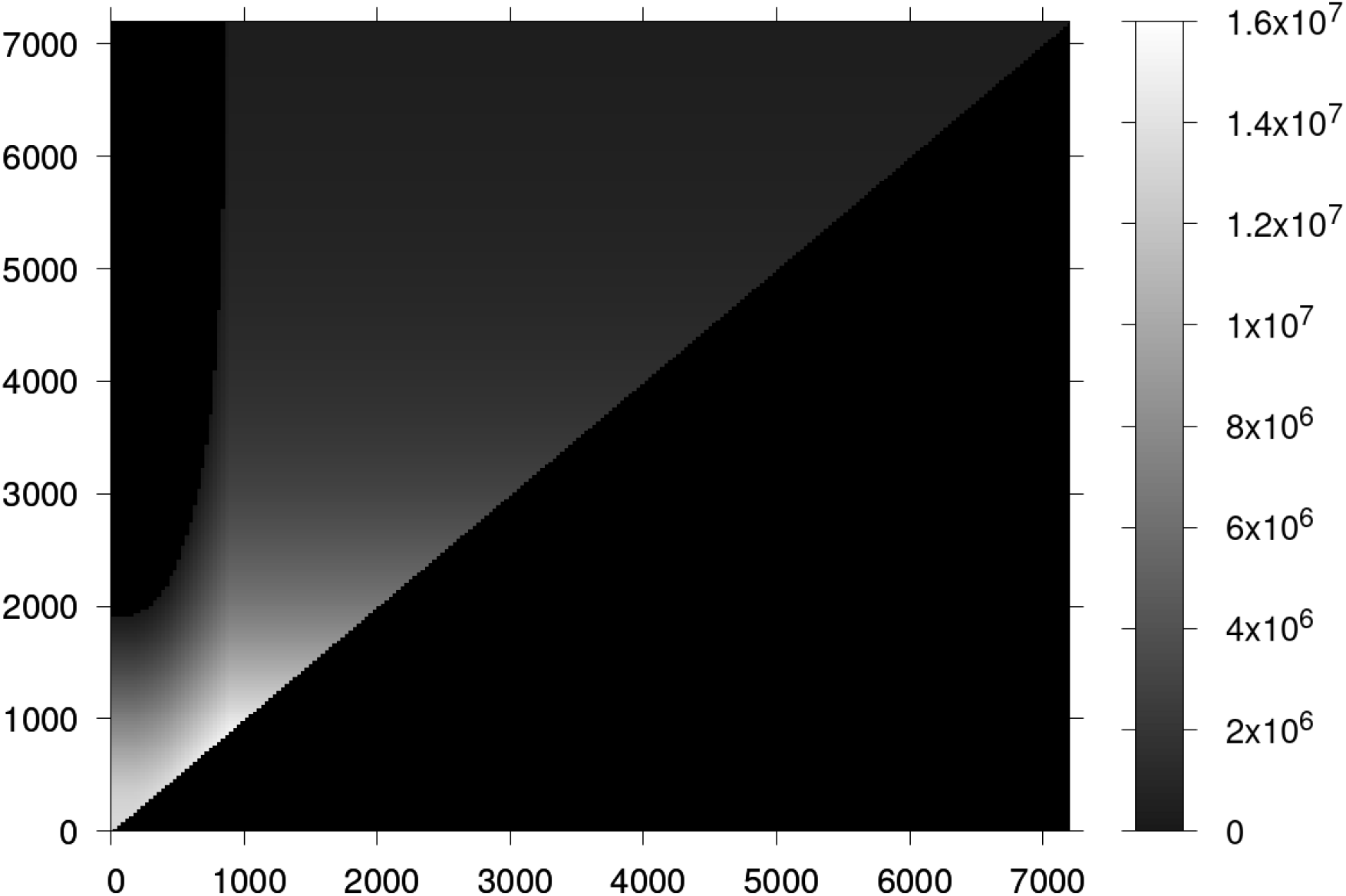
Expected value of perfect information (color scale, thousand USD) as a function of *T_v_* (x-axis) and *T_v_* + *T_c_*(y-axis). The black triangle *T_c_ <* 0 reads “not defined”.

**Figure 8:**
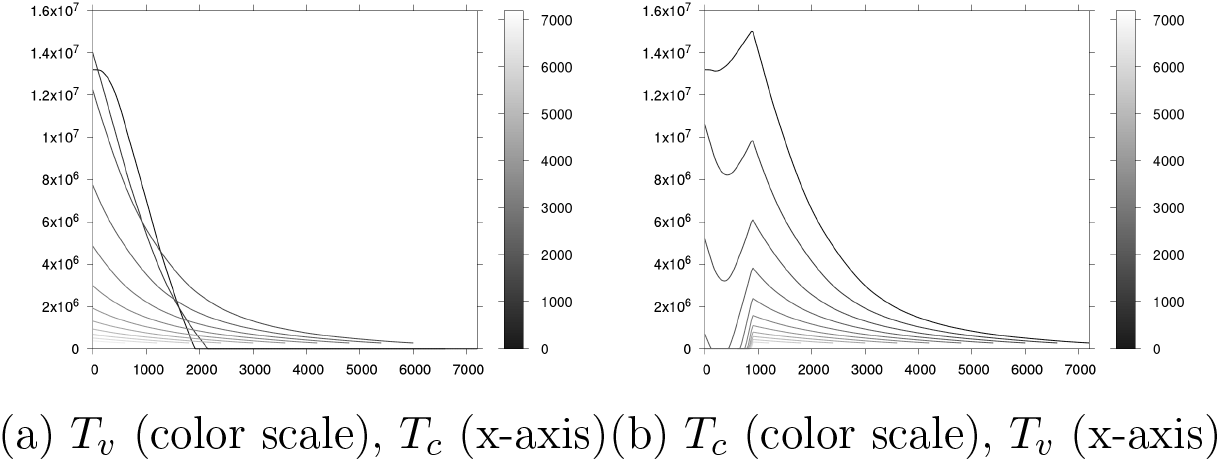
Expected value of perfect information (thousand USD) as a function of *T_c_* and *T_v_* for varying *T_v_* and *T_c_* respectively. Estimation over 10,000 parameter draws. 95% CI: see Figure App-2 in Appendix.

**Figure 9:**
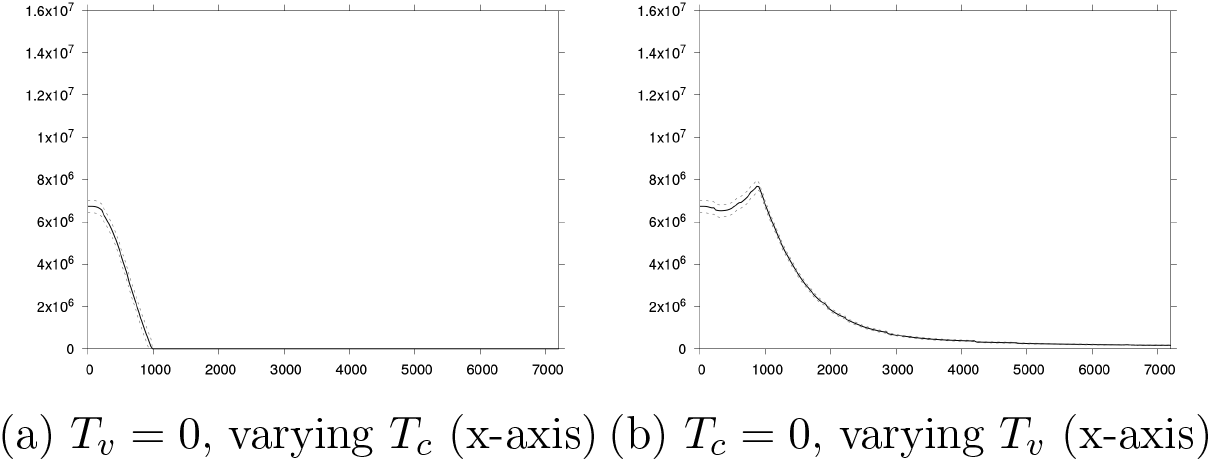
Expected value of partially perfect information (thousand USD) for vaccine parameters (*α*, *E*, *δ*, *c_v_*, and *c_a_*) as a function of *T_c_* for *T_v_* = 0 and as a function of *T_v_*for *T_c_* = 0. Estimation over 10,000 parameter draws. Dotted lines: 95% CI.

In our scenario, value of information dynamics for fixed *T_v_* as a function of *T_c_* is intuitive. From Figures 8a and 9a, we see that for any date *T_v_* at which vaccine doses are available, information has more value if it comes earlier rather than later. Value of information dynamics as a function of *T_v_* is less straightforward. We see from Figures 8b and 9b that it can be non monotonic. As expected, information becomes irrelevant for large values of *T_v_*, that is if vaccine doses are available too late – and this remains true even if clinical research can be performed quickly. For small values of *T_v_*, it can be optimal to vaccinate under uncertainty rather than wait for information if *T_c_* is too large (light grey curves in Figure 8a). However if *T_c_* is not too small and *T_v_* small enough, this effect can be reversed and the value of information increases again as *T_v_* decreases.

## 4 Conclusion

We illustrated two critical – if not defining – features of clinical research in a Disease X outbreak scenario. First, the dynamic nature of the problem of allocating resources to research. The Ebola crisis already showed that the disease transmission dynamics influences the cost of clinical trials e.g. if cases become rare or difficult to locate due to a vanishing or bursty epidemic. Here, we showed with an example that the value of information itself, that is an upper bound on the costs, depends on the date from which this information is available, but also on the date from which it can actually be used (in our scenario: the date from which vaccine doses are available). The second critical point highlighted in this article is the role of uncertainties – a central issue for Disease X preparedness. The objective of clinical research is to resolve uncertainties regarding a disease and medical countermeasures. Yet these uncertainties influence resource allocation to research. In some circumstances, making a decision under uncertainties can be optimal. While this simple idea is at the core of value of information analyses, a common approach in the fields of public health and health economics, many studies seek to design clinical trials satisfying some properties (e.g. statistical power) conditional on parameter values that would most likely be uncertain in a Disease X scenario. The reasoning in this article takes uncertainties fully into account.

Once again, all results presented in this article depend on the assumed prior beliefs or prior information. This includes parameter prior distributions, but also our assumptions regarding the transmission model structure (SIR) and the vaccine mechanism (all-or-nothing protection, no waning). Our assumptions are meant (i) to be plausible, (ii) to be concise and straightforward (we want to develop a minimal example), and (iii) to illustrate effects of interest (i.e. avoid trivial instances of the problem). We show results obtained with alternative parameter prior distributions in Appendix E. Similarly, we used an EVPI approach for concision and generality. In practice, other value of information metrics such as the expected value of sample information could be relevant, but they would require to define and discuss the details of how information is acquired (technically) for each parameter or subset of parameters, which is outside the scope of our article.

Importantly, while our study is by no way predictive of the magnitude or relevance of specific effects, our reasoning is valid in general. Moreover, our approach is based on numerical simulations of standard epidemiological models and can thus easily be adapted to pathogens or families of pathogens of interest. Replacing our illustrative SIR model by a more realistic model, modelling more realistic vaccination policies, or considering several medical countermeasures in parallel are a mere technical issues and have no bearing on the general approach. We hope that the approach developed in this article will help researchers properly take uncertainties and transmission dynamics into account when planning for Disease X clinical research and medical countermeasure development.

## Data Availability

All data produced in the present study are available upon reasonable request to the authors

## A Additional figure: transmission model

**Figure App-1:**
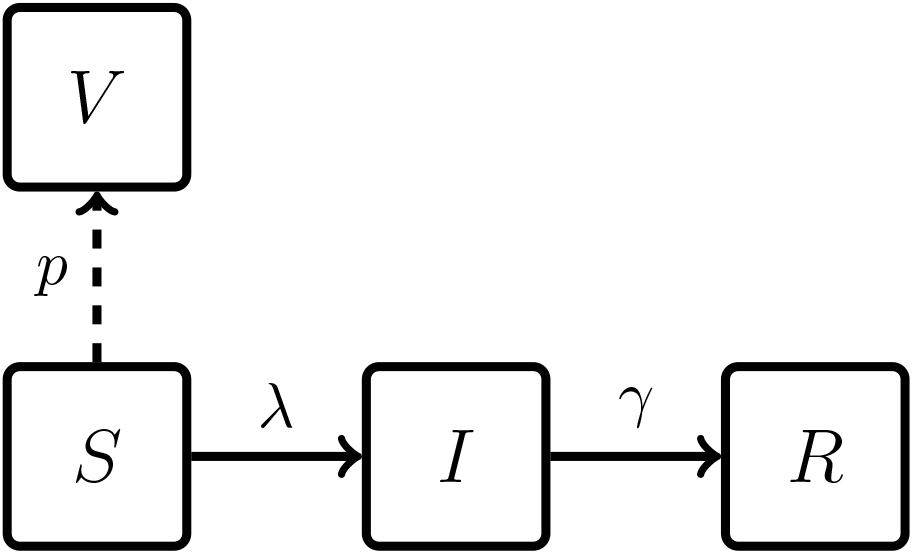
Sketch of the transmission model. Dashed: possible vaccination (instantaneous). *λ* = *γR*_0_I *N* is the force of infection.

## B Parameter prior distributions

Table App-1 shows the parameter prior distributions used in the article.

**Table App-1:** Model parameter prior distributions.

| Parameter | Distribution | Truncation |
| --- | --- | --- |
| $N$ | Constant: $10^8$ | |
| $T_v$ | Variable | |
| $T_c$ | Variable | |
| $R_0$ | Gamma distribution with mean 1.05 and standard deviation 0.2 | $[1.01; \infty]$ |
| $\gamma$ | Lognormale distribution with mean 0.1 and standard deviation 0.1 | $[0.02; \infty]$ |
| $\alpha$ | Uniform distribution | $[0, 1]$ |
| $\epsilon$ | Lognormale distribution with mean 0.003 and standard deviation 0.003 | $[0; 1]$ |
| $\delta$ | Normal distribution with mean 40 and standard deviation 20 | $[0; \infty]$ |
| $c_v$ | Normal distribution with mean 0.1 and standard deviation 0.1 | $[0; \infty]$ |
| $c_i$ | Normal distribution with mean 0.1 and standard deviation 0.03 | $[0; \infty]$ |
| $c_a$ | Normal distribution with mean 50 and standard deviation 50 | $[0; \infty]$ |

## C Additional figures: EVPI confidence intervals

In Figure App-2, we show two curves from Figure 8 with 95% confidence intervals.

**Figure App-2:**
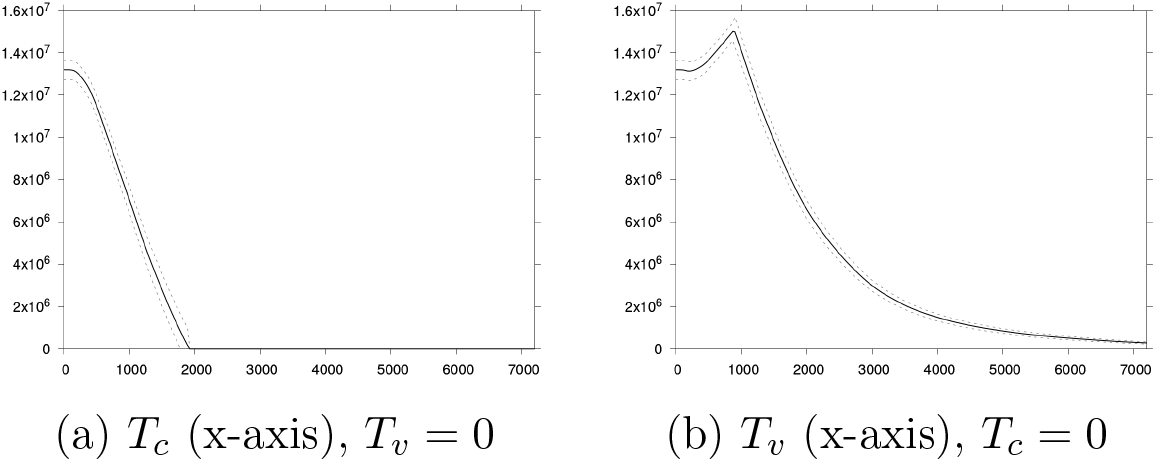
Expected value of perfect information as a function of *T_c_* and *T_v_* for *T_v_* = 0 and *T_c_* = 0 respectively. Estimation over 10,000 parameter draws. Dotted lines: 95% CI.

## D EVPPI

In Figures App-3 and App-4, we show EVPPI for different subsets of parameters as a function of *T_c_* and *T_v_*. Qualitatively, the EVPPI dynamics is similar to the EVPI dynamics. As expected, EVPPI is 0 for parameter *δ*, the average delay between vaccination and adverse event (plot not shown).

**Figure App-3:**
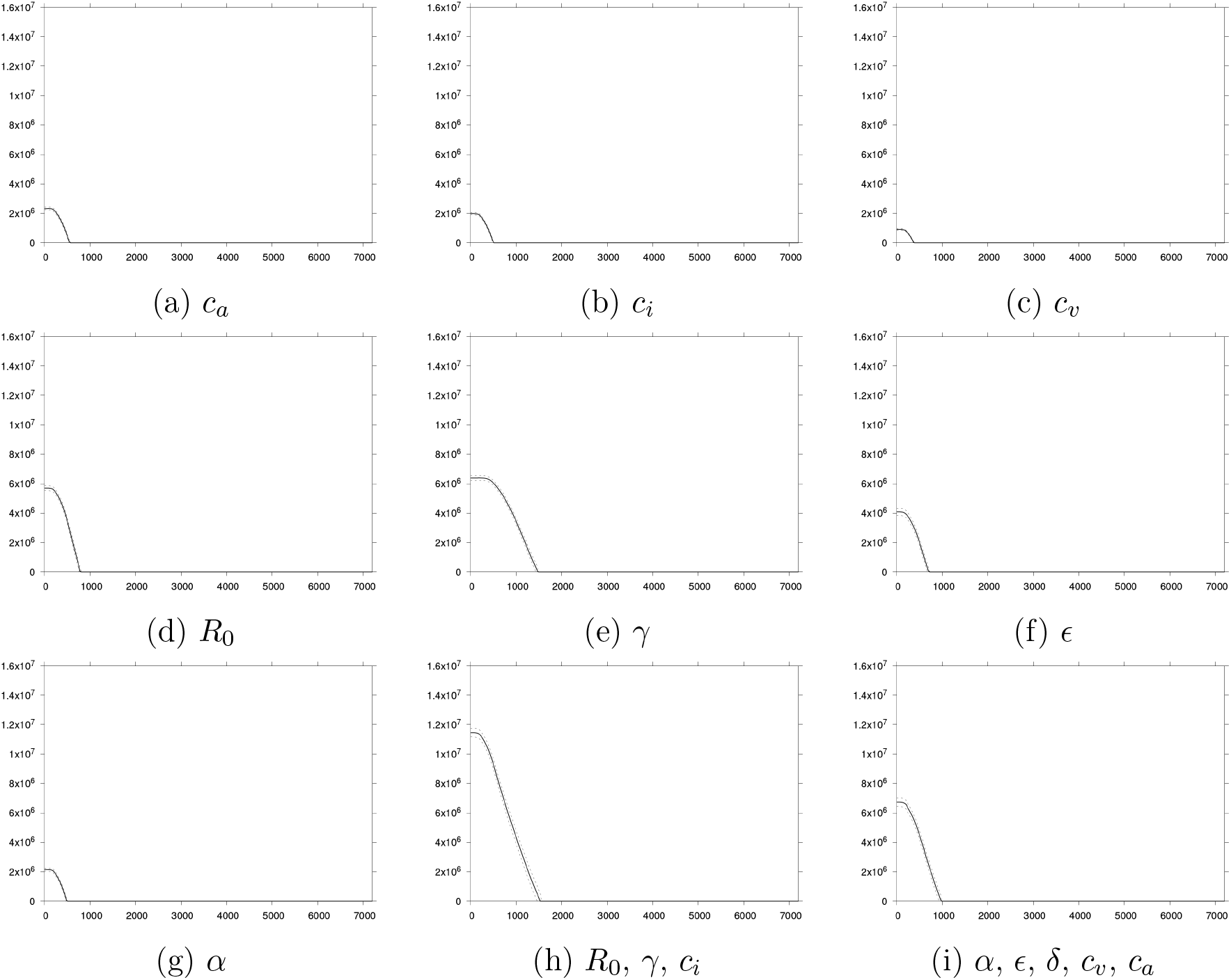
EVPPI as a function of *T_c_* in the case with *T_v_* = 0. Estimation over 10,000 parameter draws. Dotted lines: 95% CI.

**Figure App-4:**
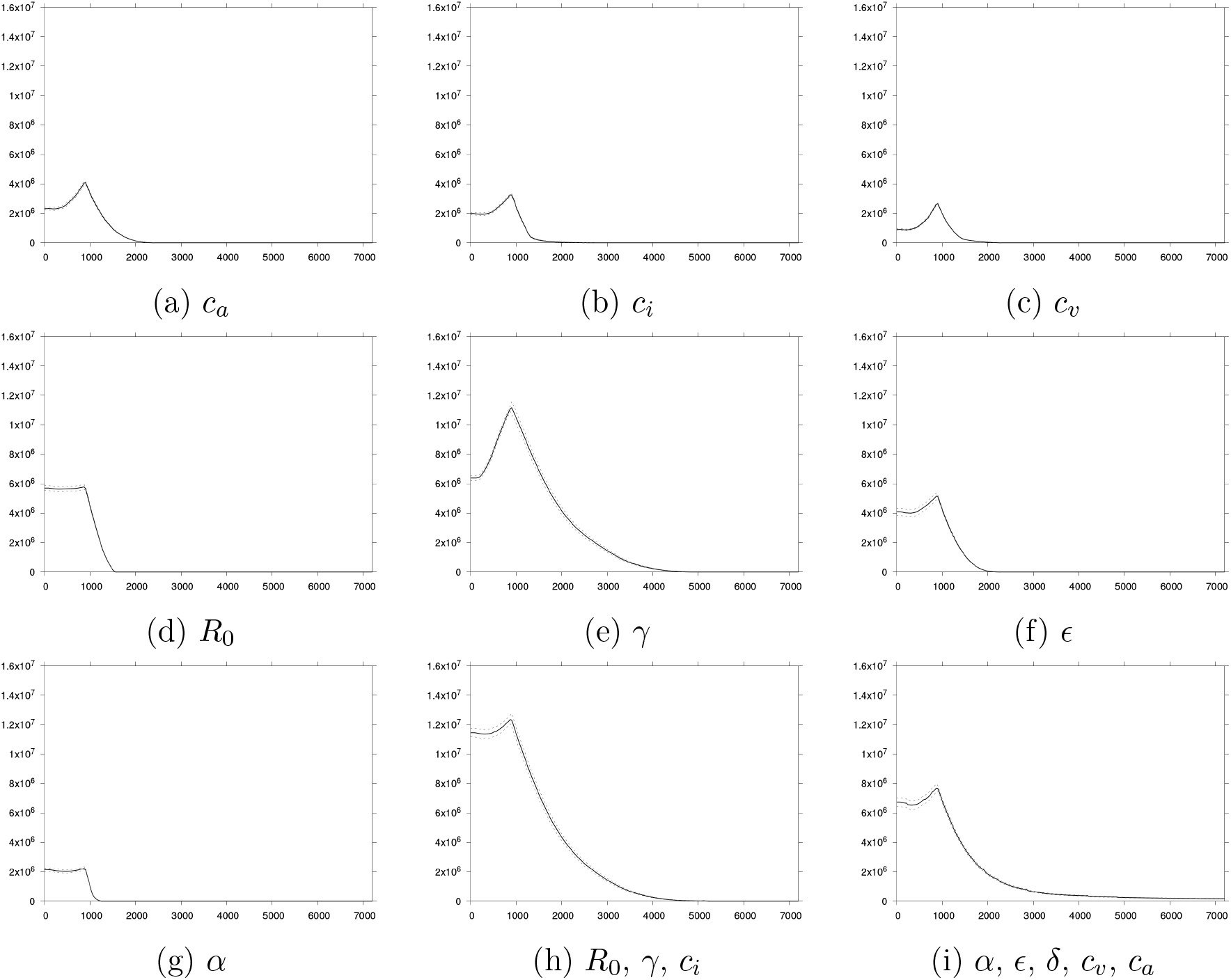
EVPPI as a function of *T_v_* in the case with *T_c_* = 0. Estimation over 10,000 parameter draws. Dotted lines: 95% CI.

**Figure App-5:**
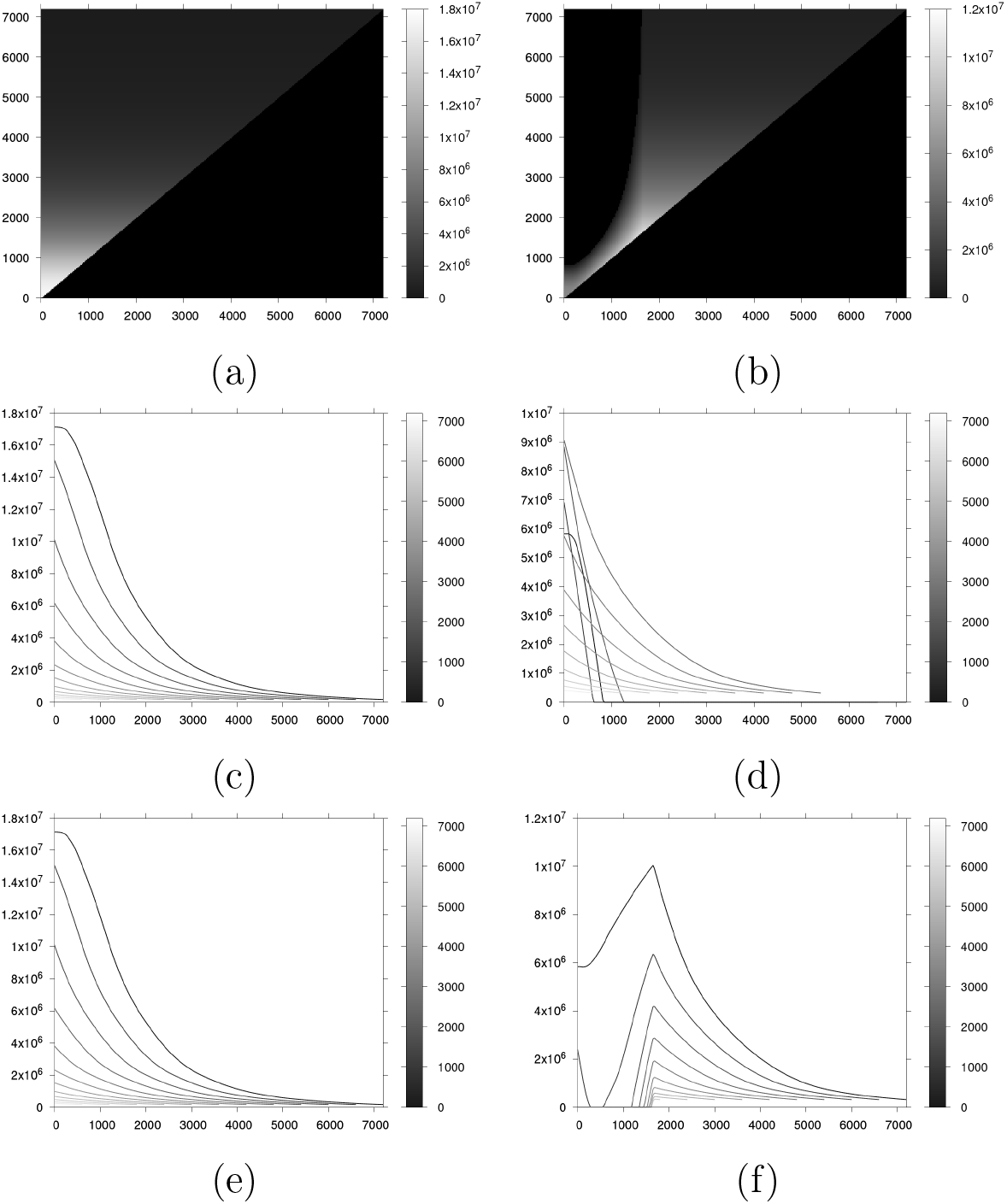
Expected value of perfect information (thousand USD) as a function of *T_c_* and *T_v_* for alternative parameter prior distributions 1 (left panels) and 2 (right panels). Figures App-5a and App-5b: EVPI (color scale) as a function of *T_v_* (x-axis) and *T_v_* + *T_c_*(y-axis). Figures App-5c and App-5d: EVPI (y-axis) as a function of *T_c_* (x-axis) *T_v_* (color scale). Figures App-5e and App-5f: EVPI (y-axis) as a function of *T_v_* (x-axis) *T_c_* (color scale). Estimations over 10,000 parameter draws.

## E Alternative prior distributions

Here we provide results for alternative prior distribution, for illustration only. The prior distributions are described in Tables App-2 and **??** and the corresponding EVPI estimates are displayed in Figure E.

**Table App-2:**
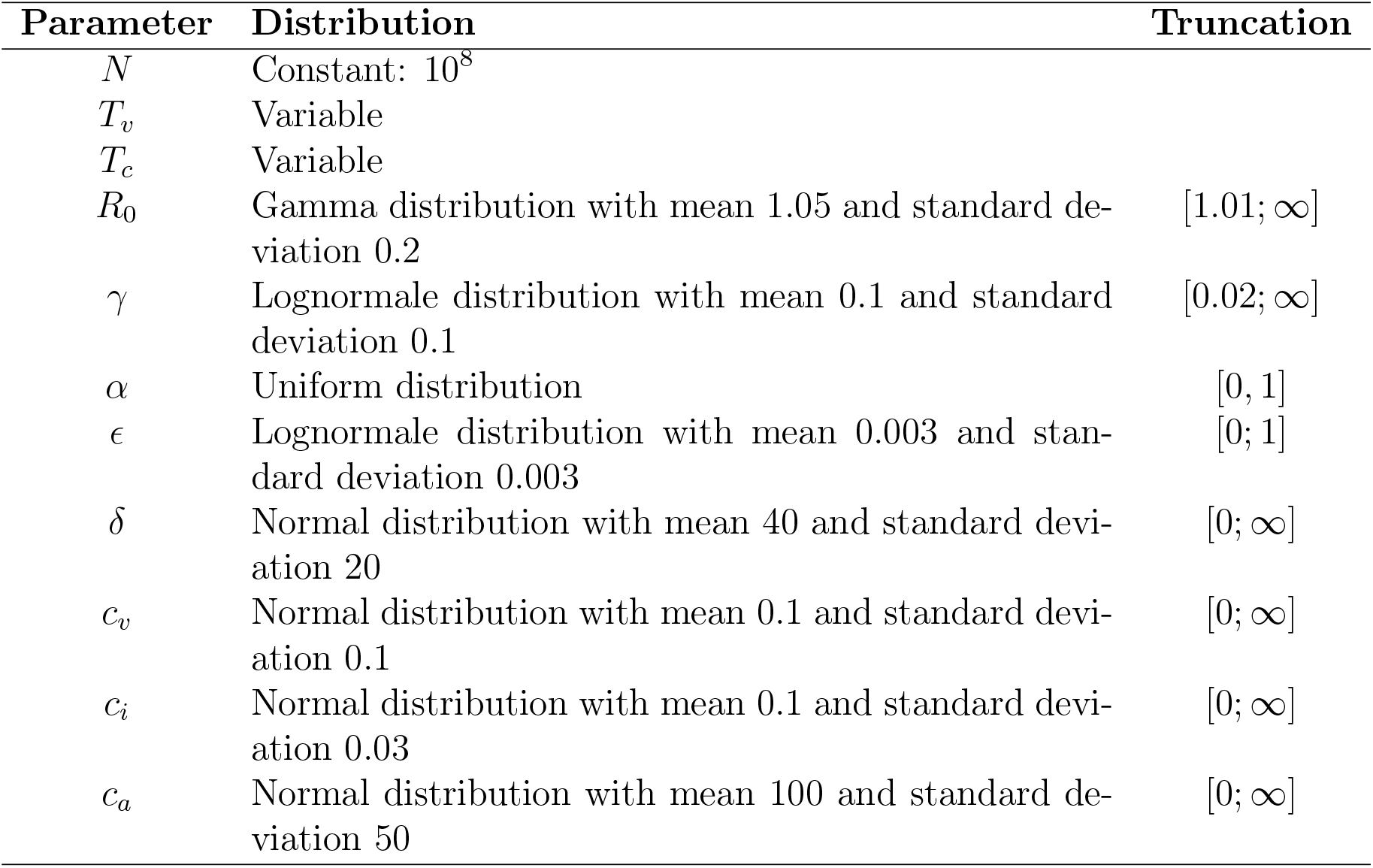
Alternative parameter prior distributions 1.

| Parameter | Distribution | Truncation |
| --- | --- | --- |
| $N$ | Constant: $10^8$ | |
| $T_v$ | Variable | |
| $T_c$ | Variable | |
| $R_0$ | Gamma distribution with mean 1.05 and standard deviation 0.2 | $[1.01; \infty]$ |
| $\gamma$ | Lognormale distribution with mean 0.1 and standard deviation 0.1 | $[0.02; \infty]$ |
| $\alpha$ | Uniform distribution | $[0, 1]$ |
| $\epsilon$ | Lognormale distribution with mean 0.003 and standard deviation 0.003 | $[0; 1]$ |
| $\delta$ | Normal distribution with mean 40 and standard deviation 20 | $[0; \infty]$ |
| $c_v$ | Normal distribution with mean 0.1 and standard deviation 0.1 | $[0; \infty]$ |
| $c_i$ | Normal distribution with mean 0.1 and standard deviation 0.03 | $[0; \infty]$ |
| $c_a$ | Normal distribution with mean 100 and standard deviation 50 | $[0; \infty]$ |

**Table App-3:**
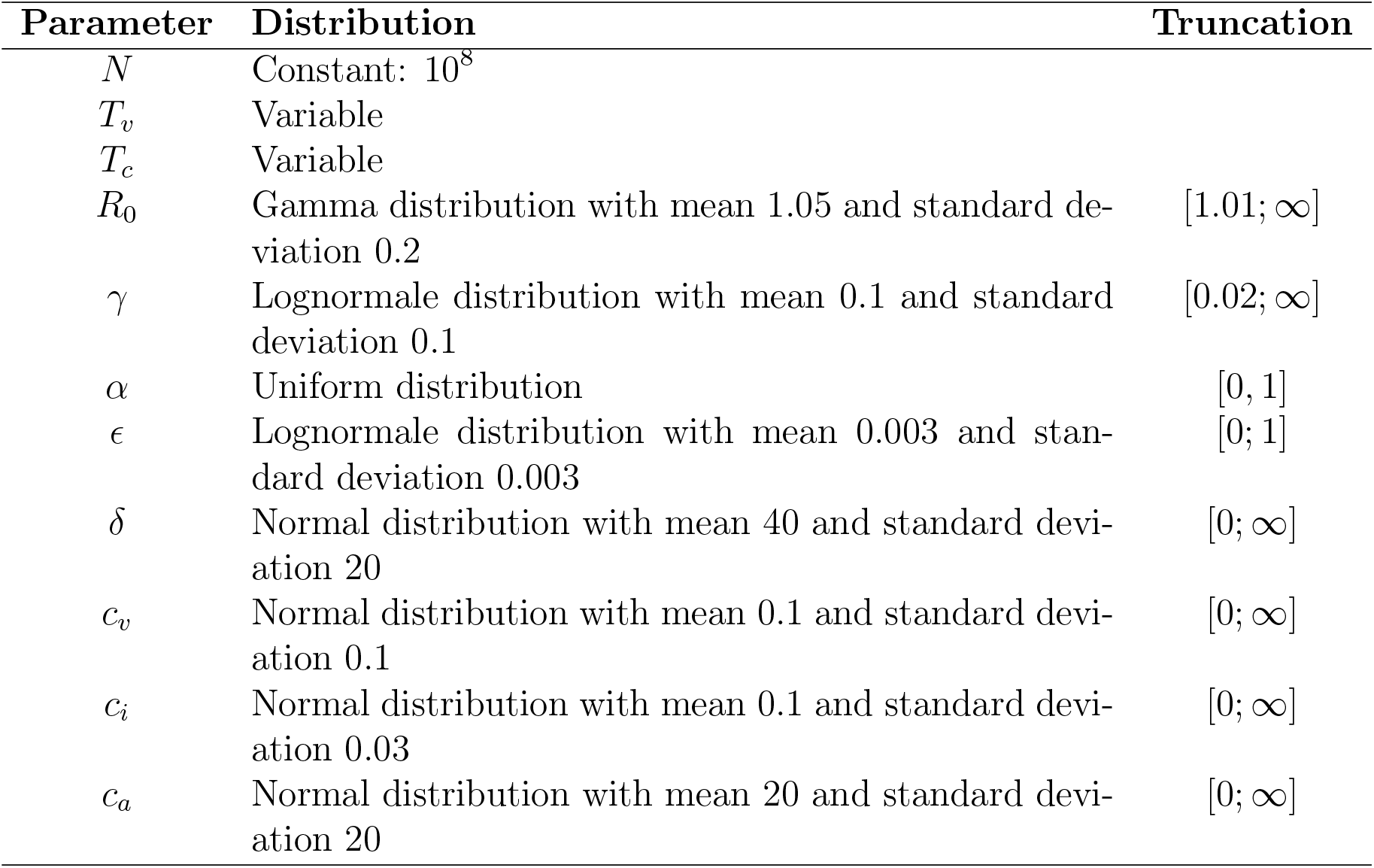
Alternative parameter prior distributions 2.

Table App-2: Alternative parameter prior distributions 1.
| Parameter | Distribution | Truncation |
| --- | --- | --- |
| $N$ | Constant: $10^8$ | |
| $T_v$ | Variable | |
| $T_c$ | Variable | |
| $R_0$ | Gamma distribution with mean 1.05 and standard deviation 0.2 | $[1.01; \infty]$ |
| $\gamma$ | Lognormale distribution with mean 0.1 and standard deviation 0.1 | $[0.02; \infty]$ |
| $\alpha$ | Uniform distribution | $[0, 1]$ |
| $\epsilon$ | Lognormale distribution with mean 0.003 and standard deviation 0.003 | $[0; 1]$ |
| $\delta$ | Normal distribution with mean 40 and standard deviation 20 | $[0; \infty]$ |
| $c_v$ | Normal distribution with mean 0.1 and standard deviation 0.1 | $[0; \infty]$ |
| $c_i$ | Normal distribution with mean 0.1 and standard deviation 0.03 | $[0; \infty]$ |
| $c_a$ | Normal distribution with mean 20 and standard deviation 20 | $[0; \infty]$ |

## Footnotes

1 The details of prior distributions are provided in Appendix B.

2 Notice that optimization techniques are not the main point of our study.

3 Notice that, for the sake of conciseness, we consider no cost for acquiring information.

